# Machine Learning Forecast of Growth in COVID-19 Confirmed Infection Cases with Non-Pharmaceutical Interventions and Cultural Dimensions: Algorithm Development and Validation

**DOI:** 10.1101/2021.01.04.21249235

**Authors:** Arnold YS Yeung, Francois Roewer-Despres, Laura Rosella, Frank Rudzicz

## Abstract

**Background:** National governments have implemented non-pharmaceutical interventions to control and mitigate against the COVID-19 pandemic. A deep understanding of these interventions is required.

**Objective:** We investigate the prediction of future daily national Confirmed Infection Growths – the percentage change in total cumulative cases across 14 days – using metrics representative of non-pharmaceutical interventions and cultural dimensions of each country.

**Methods:** We combine the OxCGRT dataset, Hofstede’s cultural dimensions, and COVID-19 daily reported infection case numbers to train and evaluate five non-time series machine learning models in predicting Confirmed Infection Growth. We use three validation methods – *in-distribution, out-of-distribution*, and *country-based cross-validation* – for evaluation, each applicable to a different use case of the models.

**Results:** Our results demonstrate high *R*^2^ values between the labels and predictions for the *in-distribution, out-of-distribution*, and *country-based cross-validation* methods (0.959, 0.513, and 0.574 respectively) using random forest and AdaBoost regression. While these models may be used to predict the Confirmed Infection Growth, the differing accuracies obtained from the three tasks suggest a strong influence of the use case.

**Conclusions:** This work provides new considerations in using machine learning techniques with non-pharmaceutical interventions and cultural dimensions data for predicting the national growth of confirmed infections of COVID-19.

## Introduction

### Background

In response to the COVID-19 pandemic, national governments have implemented non-pharmaceutical interventions (NPIs) to control and reduce the spread in their respective countries [1–4]. Indeed, early reports suggested the potential effectiveness of the implementation of NPIs to reduce the transmission of COVID-19 [2,5–9] and other infectious diseases [10–12]. Many epidemiological models forecasting future infection numbers have therefore suggested the role of NPIs in reducing infection rates [2,6,9,13], to aid with implementing national strategies and making policy decisions. In this paper, we also include the implementation of NPIs at the national level as features in predicting the national growth of the number of confirmed infection cases. Prior work has focused on the NPI variations in different regions of specific countries [2,3,5,8,14].

Various metrics may provide different perspectives and insights on the pandemic. In this study, we focus on one: Confirmed Infection Growth (CIG), which is the 14-day growth of the cumulative number of reported infection cases. Other common metrics to measure the transmission rates of an infectious disease are the basic reproduction number, *R*_0_, which measures the expected number of direct secondary infections generated by a single primary infection when the entire population is susceptible [4,15] and the effective reproduction number, *R*_*t*_ [2], which accounts for the immunity within the specified population. While such metrics are typically used by epidemiologists as measures of the transmission of an infectious disease, these metrics are dependent on estimation model structures and assumptions, making them application-specific and potentially misapplied [15]. Furthermore, the public may be less familiar with such metrics, as opposed to more practical and observable metrics, such as the absolute or relative change in cumulative reported cases.

### Related Work

Mathematical modelling of the transmission of infectious disease has been a common method to simulate infection trajectories. A common technique for epidemics is the SIR model, which separates the population into three sub-populations (susceptible, infected, and removed) and iteratively models the interaction and shift between these sub-populations, which change throughout the epidemic [16,17]. Variations of this model have since been introduced to reflect other dynamics expected of the spread of infectious diseases [18–20]. These variations of the SIR model have also been applied to the ongoing COVID-19 pandemic [21–24].

Recent studies have also used various statistical and machine learning techniques for short-term forecasting of infection rates for the COVID-19 pandemic [23,25,26], using reported transmission and mortality statistics, population geographical movement data, and media activity. Machine learning has also been used for other applications to combat the pandemic, such as in-patient monitoring and genome sequencing [27–30].

### Description of Study

The CIG because it is a verifiable metric that, due to its direct inference from the number of reported cases, may have a greater impact on the public’s perception of the magnitude of the pandemic compared to the actual transmission rate. CIG reflects the growth in the total number of reported cases within a country in 14 days relative to the total number of previously reported infections, including recoveries and mortalities. We emphasize that the reported number of infections may not necessarily be correlated with the actual transmission rate, due to factors such as different testing criteria and varying accessibility in testing over time.

To predict the CIG for individual countries, we deploy five machine learning models. We use features representing the implementation levels of NPIs and the cultural dimensions of each country. We obtain daily metrics of the implementation of NPIs at the national level from the Oxford COVID-19 Government Response Tracker (OxCGRT) dataset [1]. Although different countries may implement similar NPIs, some have suggested that cross-cultural variations across populations may lead to different perception and responses towards such NPIs [31–33]. We intend to capture any effects due to national cross-cultural differences by complementing the OxGCRT dataset with national cultural norm values from Hofstede’s cultural dimensions [34]. Our non-time series models predict the expected future national CIG using both NPI implementation and cultural norm features. While time-series deep learning models (e.g., RNNs or transformers) may also provide CIG predictions, such models generally require greater amounts of accurately labeled trajectory data and assume that past trajectory trends are readily available and representative of future trajectories. Instead, our non-time series models train on more granular data which does not necessarily need to be temporally concatenated into a trajectory. We also opt for less complex non-time series models due to indeterminacies in acquiring and verifying sufficient trajectory data, especially due to the lack of reliable data at the onset of the COVID-19 outbreak.

Our results suggest that non-time series machine learning models can predict future CIG according to multiple validation methods, depending on the user’s application. While we do not necessarily claim state-of-the-art performance for infection rate prediction given the rapidly growing amount of parallel work in this area, to the best of our knowledge, our work is the first to use machine learning techniques to predict the change in national cumulative numbers of reported COVID-19 infections by combining NPI implementation features with national cultural features. Our implementation uses publicly available data retrieved from the Internet and relies on the open-sourced Python libraries Pandas [35] and Scikit-Learn [36].

## Methods

### Data and Pre-Processing

Candidate features at the national level are extracted from three datasets for input into our machine learning models: NPIs, cultural dimensions, and current COVID-19 confirmed case numbers.

The Oxford COVID-19 Government Response Tracker (OxCGRT) provides daily level metrics of the NPIs implemented by countries [1]. This dataset categorizes NPIs into 17 categories, each with either an ordinal policy level metric ranging from (not implemented) to 3 (strictly enforced) or a continuous metric, representing a monetary amount (e.g., research funding). We limit our candidate features to the 13 ordinal policy categories, as well as 4 computed indices, which represent the implementation of different policy types taken by governments, based on the implemented NPIs. This dataset contains data starting from 1 January 2020.

To represent cultural differences across populations of different countries, the 2015 edition of Hofstede’s cultural dimensions [37] are tagged to each country. While rarely used in epidemiology studies, these dimensions have been used frequently in international marketing studies and cross-cultural research as indicators of the cultural values of national populations [38]. Because the 2015 edition of this dataset groups certain geographically neighboring countries together (e.g., Ivory Coast, Burkina Faso, Ghana, etc. into Africa West), we tag all subgroup countries with the dimension values of their group. While we recognize this is far from ideal and will likely lead to some degree of inaccurate approximation in these subgroup countries, we perform this pre-processing step to include those countries in our study. The dimension values for each country are constant across all samples. Six cultural dimensions are presented for each country/region [39]:

- *Power distance index:* The establishment of hierarchies in society and organizations, and the extent to which lower hierarchical members accept inequality in power.
- *Individualism v. collectivism:* The degree to which individuals are not integrated into societal groups (e.g., individual, immediate family (*individualistic*) v. extended families (*collectivistic*))
- *Uncertainty avoidance:* Society’s tendency to avoid uncertainty and ambiguity through use of the societal disapproval, behavioral rules, laws, etc.
- *Masculinity v. femininity:* Societal preference towards assertiveness, competitiveness, and division in gender roles (*masculinity*), compared to caring, sympathy, and similarity in gender roles (*femininity*)
- *Long-term v. short-term orientation:* Societal values towards tradition, stability, and steadfastness (*short-term*) v. adaptability, perseverance, and pragmatism (*long-term*)
- *Indulgence v. restraint:* The degree of freedom available to individuals for fulfilling personal desires by social norms (e.g., free gratification (*indulgence*) v. controlled gratification (*restraint*))

We extract the daily number of confirmed cases, *n*_*t*_, for each country from the COVID-19 Data Repository by the Center for Systems Science and Engineering (CSSE) at Johns Hopkins University [40]. We use a rolling average of the previous 5-day window to smooth fluctuations in *n*_*t*_, which may be caused by various factors, such as inaccurate case reporting, no release of confirmed case numbers (e.g., on weekends and holidays), and sudden infection outbreaks. We refer to the smoothed daily number of confirmed cases for date as 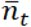.

We compute the Confirmed Infection Growth (CIG) for a specified date, *τ*, as:

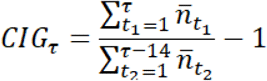

The CIG represents the expected number of new confirmed cases from date *τ* − 13 to date as a percentage of the total number of confirmed infection cases up to date *τ* − 14.

Our goal is to predict the CIG 14 days in advance (i.e., *CIG*_*τ* + 14_) given information from the current date *τ* for each country. Candidate features available include all ordinal policy metrics and the 4 computed indices from OxCGRT, the six dimension values from Hofstede’s cultural dimensions, the CIG of the current date *CIG*_*τ*_, and the smoothed cumulative number of confirmed cases 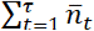, for a total of 25 feature candidates. Neither the date nor any other temporal features are included.

We trim samples with fewer than 10 cumulative confirmed infection cases and with the highest 2.5 % and the lowest 2.5 % of *CIG*_*τ* + 14_ to remove outliers in the data. Because the lowest 2.5 % of *CIG*_*τ* + 14_ are all 0.0%, we remove the samples with *CIG*_*τ* + 14_=0.0% by ascending date.

Our data ranges from 1 April 2020 to 30 September 2020 inclusively. We exclude all countries in our combined dataset that have missing feature values. In total, our combined dataset and our experiments apply to 114 countries.

### Feature Selection and Processing

We select features to input into our machine learning models from our candidate feature pool using mutual information [41]. Mutual information is a measure of the dependency between the individual feature (i.e., independent variable) and the label (i.e., dependent variable) and captures both linear and non-linear dependencies. However, mutual information does not capture multivariate dependencies nor indicates collinearity between features. To include both linear and non-linear dependencies, features are selected if they achieve a substantially non-zero mutual information, i.e., greater than 0.10. Feature selection is conducted prior to training with the training set in all validation methods. Similar feature filtering and selection techniques have been used in other machine learning applications [36,42]. The candidate features considered for input and their respective mutual information are listed in Table 1 for the in-distribution and out-of-distribution validation methods. Mutual information is also computed for each of the ten folds of the cross-validation method.

**Table 1.** Mutual information of candidate features for the in-distribution and out-of-distribution validation methods. In the cross-validation method, the ten folds have varying mutual information. Selected features are indicated with (*) for the in-distribution method and with (^†^) for the out-of-distribution method.

| Candidate Features | In-Distribution | Out-of-Distribution |
| --- | --- | --- |
| *Non-Pharmaceutical Interventions* |  |  |
| School closing<sup>*†</sup> | 0.184 | 0.205 |
| Workplace closing<sup>†</sup> | 0.098 | 0.127 |
| Cancel public events<sup>†</sup> | 0.089 | 0.127 |
| Restrictions on gatherings<sup>*†</sup> | 0.107 | 0.112 |
| Close public transport<sup>†</sup> | 0.094 | 0.124 |
| Stay at home requirements<sup>*†</sup> | 0.139 | 0.163 |
| Restrictions on internal movement<sup>*†</sup> | 0.126 | 0.146 |
| International travel controls | 0.099 | 0.099 |
| Income support<sup>†</sup> | 0.095 | 0.110 |
| Debt/contract relief | 0.043 | 0.053 |
| Public information campaigns | 0.020 | 0.023 |
| Testing policy | 0.056 | 0.064 |
| Contact tracing | 0.030 | 0.038 |
| Stringency Index<sup>*†</sup> | 0.638 | 0.668 |
| Government Response Index<sup>*†</sup> | 0.634 | 0.641 |
| Containment Health Index<sup>*†</sup> | 0.621 | 0.655 |
| Economic Support Index<sup>*†</sup> | 0.119 | 0.124 |
| *Current Infection Numbers* |  |  |
| Current cumulative number of confirmed cases: $\sum_{t=1}^{\tau} \bar{n}_t$ <sup>*†</sup> | 0.517 | 0.557 |
| $CIG_{\tau}^{*\dagger}$ | 0.866 | 0.798 |
| *Hofstede's Cultural Dimensions* |  |  |
| Power distance<sup>*†</sup> | 0.288 | 0.342 |
| Individualism<sup>*†</sup> | 0.309 | 0.355 |
| Masculinity<sup>*†</sup> | 0.310 | 0.372 |
| Uncertainty avoidance<sup>*†</sup> | 0.314 | 0.370 |
| Long-term orientation<sup>*†</sup> | 0.461 | 0.535 |
| Indulgence<sup>*†</sup> | 0.456 | 0.529 |

All selected features are then normalized to the range [0, 1]using standard min-max normalization.

### Model Training and Validation

We train the machine learning models with the combinations of hyperparameters listed in Table 2 [36,43–46]. We optimize the models using the mean squared error (MSE) criterion and select the model hyperparameters with the lowest mean absolute error (MAE) as the optimal configuration of a model. While the MSE heavily penalizes large residual errors disproportionately, the MAE provides an absolute mean of all residual errors [47]. The MAE of the training data acts as a measure of the goodness-of-fit of the model, while the MAE of the validation and testing data acts as a measure of the predictive performance [48].

**Table 2.**
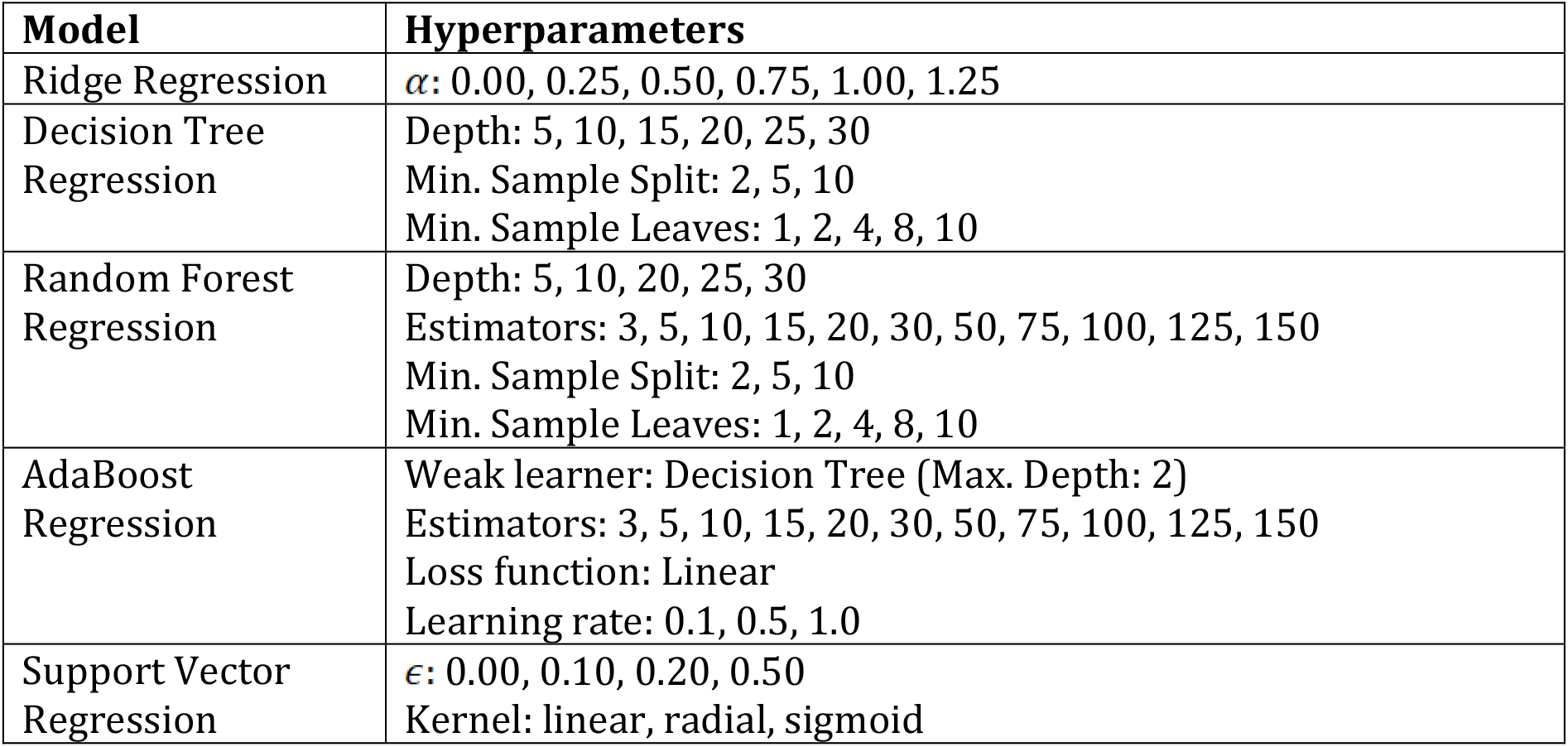
Machine learning models and hyperparameter combinations.

To validate in-distribution and out-of-distribution, we split our samples into 70-15-15 training-validation-test sets. For cross-validation [49,50], we split our samples into 10 folds (i.e., 90-10). These three methods of validation each represent a different definition of performance for the machine learning models.

#### In-distribution validation

We randomly split the samples into training, validation, and test sets. Consequently, the models are trained from samples distributed across the entire date range available in our data. This is critical - it is generally expected that model performance is best when training and test data are drawn from the same distribution. Because the COVID-19 infection numbers are naturally time series, this method ensures that validation and test samples are indeed from the same distribution as training samples. Because samples are disassociated from their dates and all other known temporal features, the prediction of the validation and test samples using the training samples are unordered. This method may be applicable to use cases where the date-to-predict is expected to be in a similar distribution as the training samples, such as predicting *CIG*_*τ* + 14_when data up to the current date is available.

#### Out-of-distribution validation

While the in-distribution method may ensure that the training, validation, and test data are all sampled from the same distribution, it may not necessarily be the most practical. Generally, the goal of long-term infection rate forecasting is to anticipate for future infection rates and should not be represented as an in-distribution task, where we have trained with data from near or later than the date-to-predict. Therefore, we also validate the performance of our models by training on the earliest 70% of samples. The validation and test sets are then randomly split between the remaining 30% of samples. This setup ensures that all training samples occurred earlier than validation and testing samples and no temporal features (known or hidden) are leaked. However, due to the changing environment related to COVID-19 infections (e.g., introduction of new NPIs, seasonal changes, new research), the validation and testing distributions is likely different from that of the training set. This method may be applicable for use cases where the date-to-predict is in the far future and not all data up to 14 days prior to the date-to-predict are available.

#### Country-based cross-validation

As a compromise between the above two methods, we also use a cross-validation method by splitting the available countries into ten folds. The aim is to evaluate validation samples from the same date range as training samples, but not the same country trajectory. That is, only data from countries not in the validation set are included in the training set. Although the samples from the training and validation sets are therefore sampled from different distributions (i.e., different countries), we anticipate that features from Hofstede’s cultural dimensions [34] may assist in identifying similar characteristics between countries, thus reducing the disparity between the training and validation distributions. This method may be applicable in predicting the CIG of countries where associated previous data is unavailable or unreliable.

## Results

### Feature Selection

For both the interpolation and extrapolation training sets, we observe that most candidate features meet our requirement of a non-zero mutual information (≥ 0.10) (see Table 1).

In both training sets, the candidate features that do *not* meet the requirements are international travel control (0.099, 0.099), debt/contract relief (0.043, 0.053), public information campaigns (0.020, 0.023), testing policy (0.056, 0.064), and contact tracing (0.030, 0.038). Additional candidate features which do not meet the requirements for interpolation training set are workplace closing (0.098) and cancel public events (0.089). Overall, the in-distribution and out-of-distribution datasets contain 17 and 20 features, respectively.

*CIG*_*τ*_ has the highest mutual information out of all features, suggesting similarities between the feature *CIG*_*τ*_ and the label *CIG*_*τ* + 14_. Further analysis shows a correlation of *r* = 0.309 between *CIG*_*τ*_ and *CIG*_*τ* + 14_. This may be due to similar trends in the CIG when the implementation of NPIs is consistent within a 14-day period.

### Comparison of Machine Learning Models

Out of all available configurations (i.e. hyperparameter combinations) of each model, we select the model configuration with the lowest validation error and compute the test error. The parameters for these selected models are listed in Table 3. The mean training, validation, and test errors are included in Table 4, Table 5, and Table 6, respectively, for the in-distribution, out-of-distribution, and cross-validation methods. We also include the median percent error [51], which is the percentage difference of the prediction *f*(*x*^(*i*)^) and the label *y*^(*i*)^ for each instance {*x*^(*i*)^, *y*^(*i*)^}, computed as:

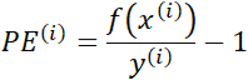

**Table 3.** Hyperparameters of the optimal configuration (lowest validation MAE) for each model for each validation method.

|  | **In-Distribution** | **Out-of-Distribution** | **Cross-Validation** |
| --- | --- | --- | --- |
| Ridge Regression | $\alpha$: 0.00 | $\alpha$: 0.25 | $\alpha$: 0.00 |
| Decision Tree Regression | Depth: 25<br>Min. Sample Split: 2<br>Min. Sample Leaves: 1 | Depth: 10<br>Min. Sample Split: 5<br>Min. Sample Leaves: 1 | Depth: 5<br>Min. Sample Split: 2<br>Min. Sample Leaves: 4 |
| Random Forest Regression | Depth: 30<br>Estimators: 150<br>Min. Sample Split: 2<br>Min. Sample Leaves: 1 | Depth: 15<br>Estimators: 10<br>Min. Sample Split: 2<br>Min. Sample Leaves: 10 | Depth: 15<br>Estimators: 125<br>Min. Sample Split: 2<br>Min. Sample Leaves: 10 |
| AdaBoost Regression | Estimators: 5<br>Learning rate: 0.1 | Estimators: 5<br>Learning rate: 1.0 | Estimators: 3<br>Learning rate: 0.1 |
| Support Vector Regression | $\epsilon$: 0.00<br>Kernel: radial | $\epsilon$: 0.00<br>Kernel: linear | $\epsilon$: 0.00<br>Kernel: linear |

**Table 4.**
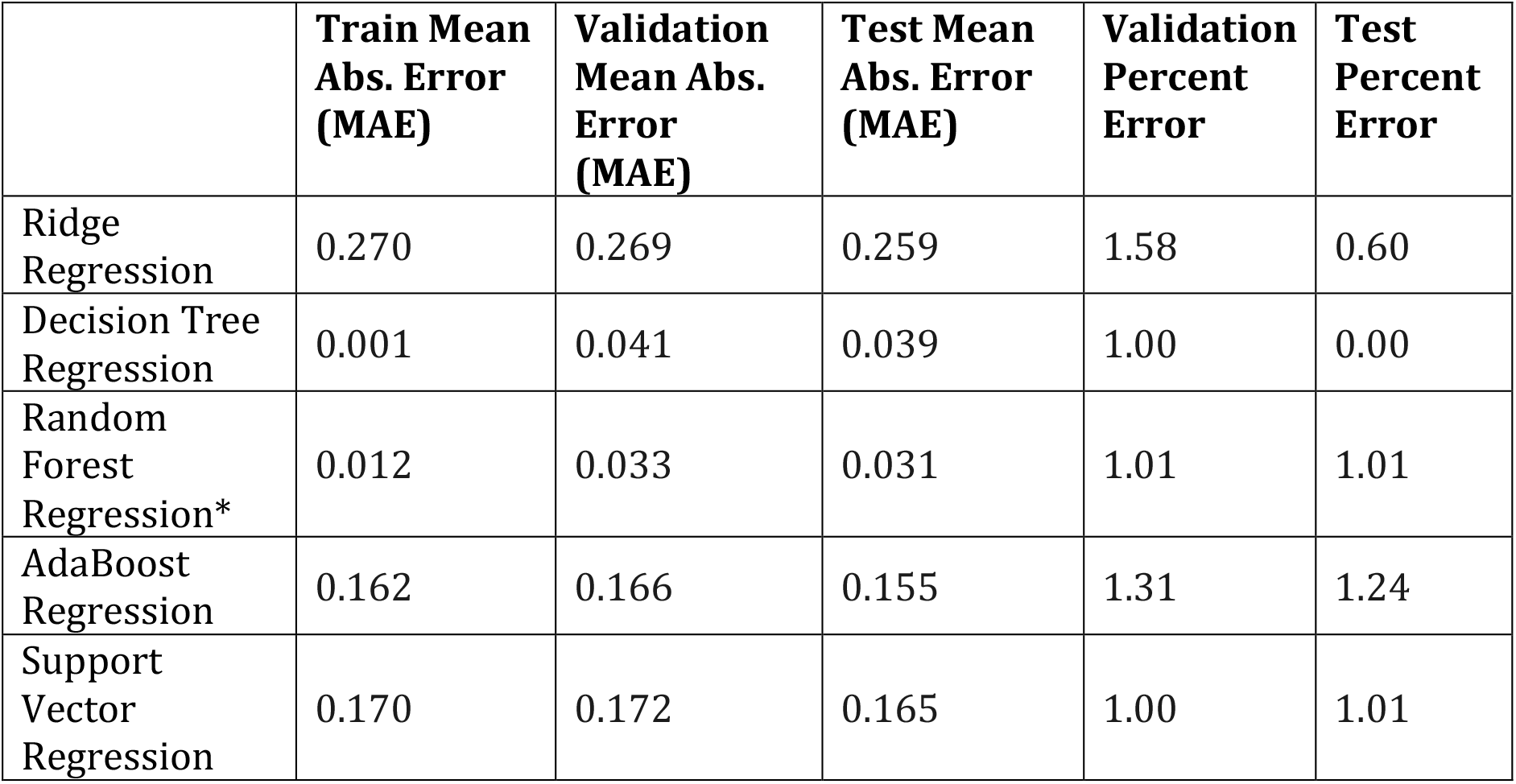
Optimal mean absolute error and median percent error for in-distribution validation method. The model indicated with the lowest test MAE is indicated with (*).

|  | **Train Mean Abs. Error (MAE)** | **Validation Mean Abs. Error (MAE)** | **Test Mean Abs. Error (MAE)** | **Validation Percent Error** | **Test Percent Error** |
| --- | --- | --- | --- | --- | --- |
| Ridge Regression | 0.270 | 0.269 | 0.259 | 1.58 | 0.60 |
| Decision Tree Regression | 0.001 | 0.041 | 0.039 | 1.00 | 0.00 |
| Random Forest Regression* | 0.012 | 0.033 | 0.031 | 1.01 | 1.01 |
| AdaBoost Regression | 0.162 | 0.166 | 0.155 | 1.31 | 1.24 |
| Support Vector Regression | 0.170 | 0.172 | 0.165 | 1.00 | 1.01 |

**Table 5.** Optimal mean absolute error and median percent error for out-of-distribution validation method. The model indicated with the lowest test MAE is indicated with (*).

|  | **Train Mean Abs. Error (MAE)** | **Validation Mean Abs. Error (MAE)** | **Test Mean Abs. Error (MAE)** | **Validation Percent Error** | **Test Percent Error** |
| --- | --- | --- | --- | --- | --- |
| Ridge Regression | 0.296 | 0.240 | 0.247 | 2.26 | 1.22 |
| Decision Tree Regression | 0.117 | 0.109 | 0.114 | 1.15 | 0.12 |
| Random Forest Regression | 0.098 | 0.098 | 0.105 | 1.45 | 0.44 |
| AdaBoost Regression* | 0.207 | 0.081 | 0.089 | 1.40 | 0.39 |
| Support Vector Regression | 0.268 | 0.167 | 0.176 | 1.66 | 0.60 |

**Table 6.** Optimal mean absolute error and median percent error for cross-validation method. (Validation error is equivalent to test error for cross-validation.) The model indicated with the lowest test MAE is indicated with (*).

|  | **Train Mean Abs. Error (MAE)** | **Validation Mean Abs. Error (MAE)** | **Validation Percent Error** |
| --- | --- | --- | --- |
| Ridge Regression | 0.262 | 0.275 | 0.62 |
| Decision Tree Regression | 0.181 | 0.207 | 0.28 |
| Random Forest Regression | 0.073 | 0.175 | 0.40 |
| AdaBoost Regression* | 0.164 | 0.167 | 0.27 |
| Support Vector Regression | 0.230 | 0.240 | 0.03 |

We observe that random forest regression has the lowest mean test error in the interpolation method (0.031) and AdaBoost regression has the lowest mean test errors in the extrapolation and cross-validation methods (0.089 and 0.167 respectively) (see Table 4, Table 5, and Table 6). For all models aside from ridge regression, the in-distribution method has the lowest mean test errors and the lowest median percent error.

### Analysis of Best Performing Models

Intercepts near 0.0 and slopes near 1.0 are the linear calibration measures that would indicate a perfect calibration relationship between the predictions and the labels [48]. For the optimal models in all validation methods, we observe slopes close to 1.0 and intercepts close to 0.0 (see Table 7). Due to the large sample sizes, statistical significance testing indicates several slopes and intercepts as significantly different from 1.0 and 0.0, respectively. However, the small mean differences (standardized to the standard deviation, i.e., z-score) indicate these differences have no practical significance. High correlations and *R*^2^ values between the predictions and labels are observed in all three validation methods (see Figure 1, Figure 2, and Figure 3).

**Table 7.** Linear calibration measures of the models with the lowest test MAE for each validation method.

|  | **In-Distribution** | **Out-of-Distribution** | **Cross-Validation** |
| --- | --- | --- | --- |
| Test Sample Size, $N$ | 2847 | 2811 | 19,669 |
| Model | Random Forest | AdaBoost | AdaBoost |
| Correlation, $r$ | 0.979 | 0.716 | 0.758 |
| Slope ± SE | $1.037 \pm 0.004$ | $0.986 \pm 0.018$ | $0.968 \pm 0.006$ |
| Slope standardized mean difference (z-score) from 1 | 0.176 | -0.015 | -0.039 |
| Slope $P$ value (mean of 1) | < 0.001 | 0.427 | < 0.001 |
| Intercept ± SE | $-0.013 \pm 0.002$ | $-0.011 \pm 0.004$ | $0.006 \pm 0.003$ |
| Intercept standardized mean difference (z-score) from 0 | -0.119 | -0.044 | 0.014 |
| Intercept $P$ value (mean of 0) | <0.001 | 0.019 | 0.059 |
| $R^2$ value | 0.959 | 0.513 | 0.574 |

**Figure 1.**
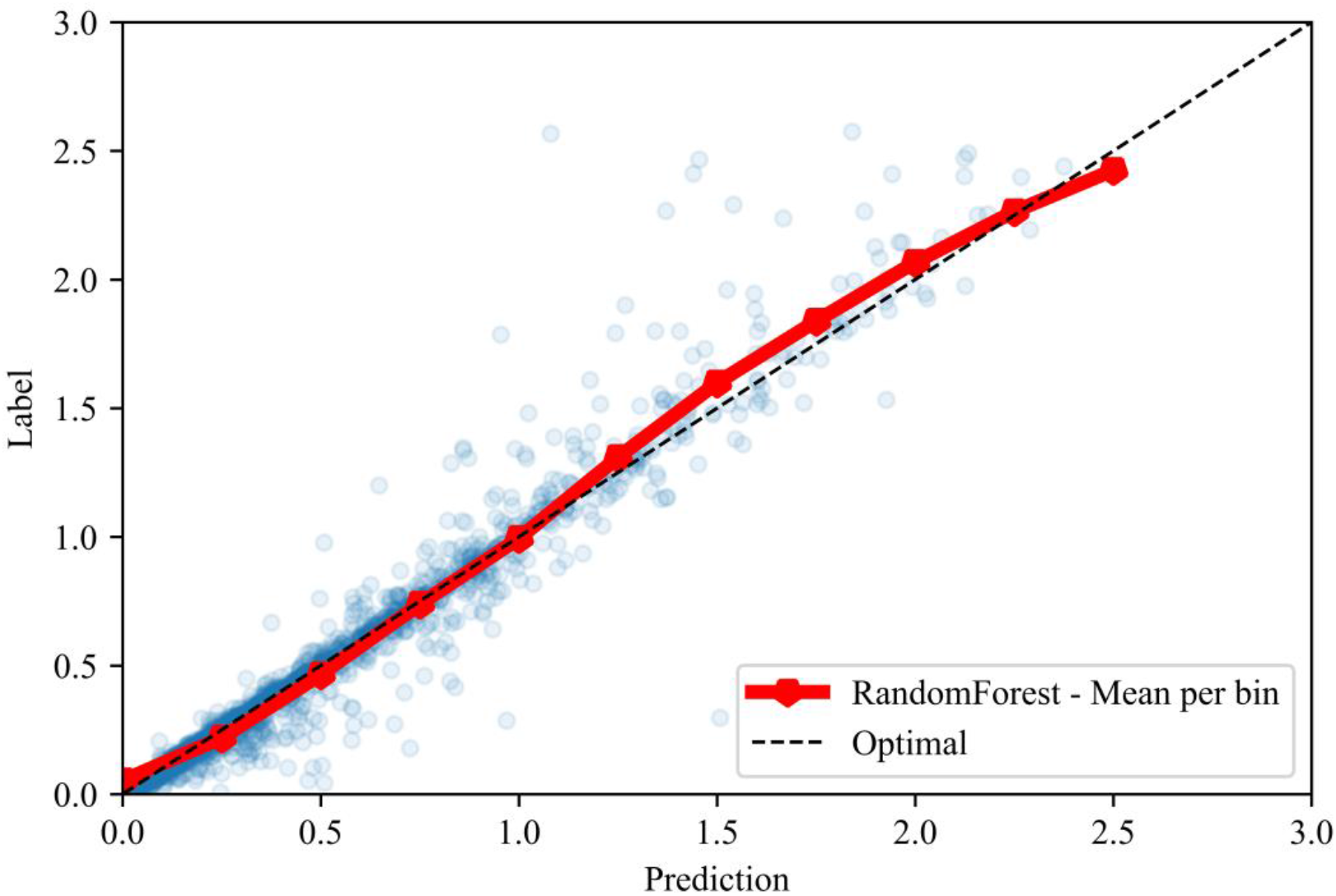
Calibration plot between labels and predictions for the interpolation validation method, with the means of each prediction bin of size 0.25.

**Figure 2.**
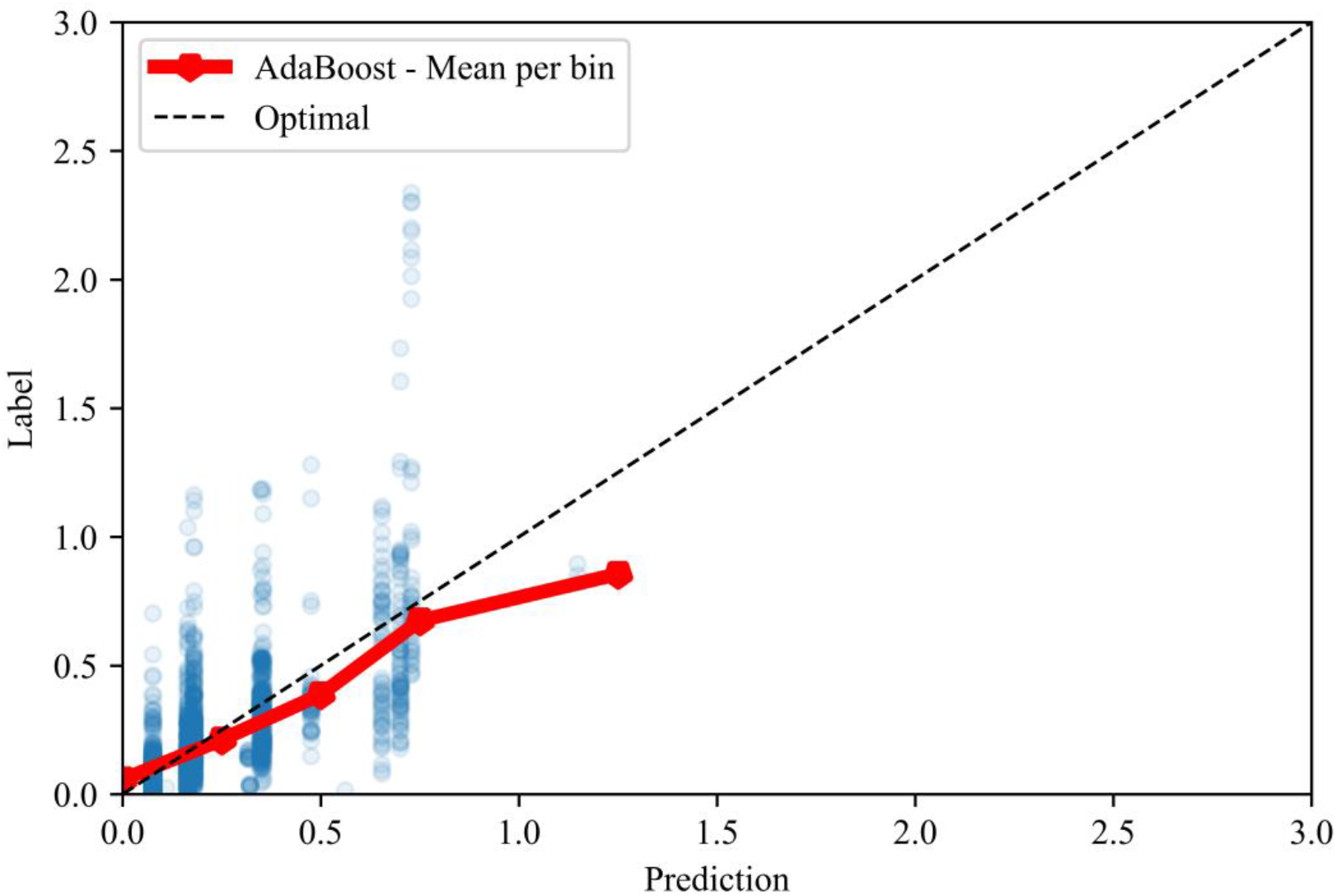
Calibration plot between labels and predictions for the extrapolation validation method, with the means of each prediction bin of size 0.25.

**Figure 3.**
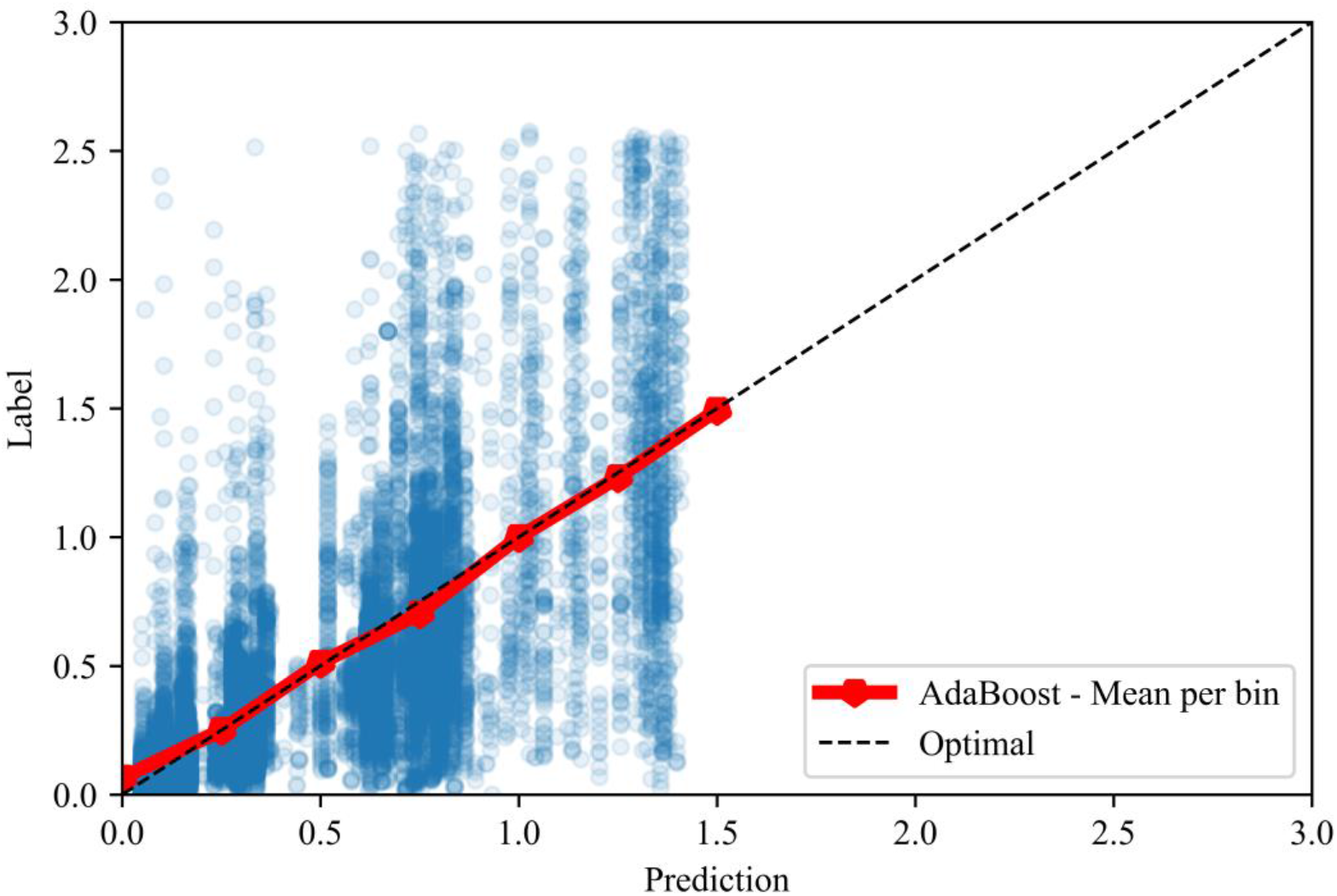
Calibration plot between labels and predictions for the cross-validation method, with the means of each prediction bin of size 0.25.

From observing the distributions of predictions and labels in all three validation methods (see Figure 4, Figure 5, and Figure 6), we see that the distributions of predictions and labels in the in-distribution method are similar. In the cross-validation method, predictions are slightly higher than the labels for the label range from 0.0 to 1.0, showing overestimation of the CIG within this range.

**Figure 4.**
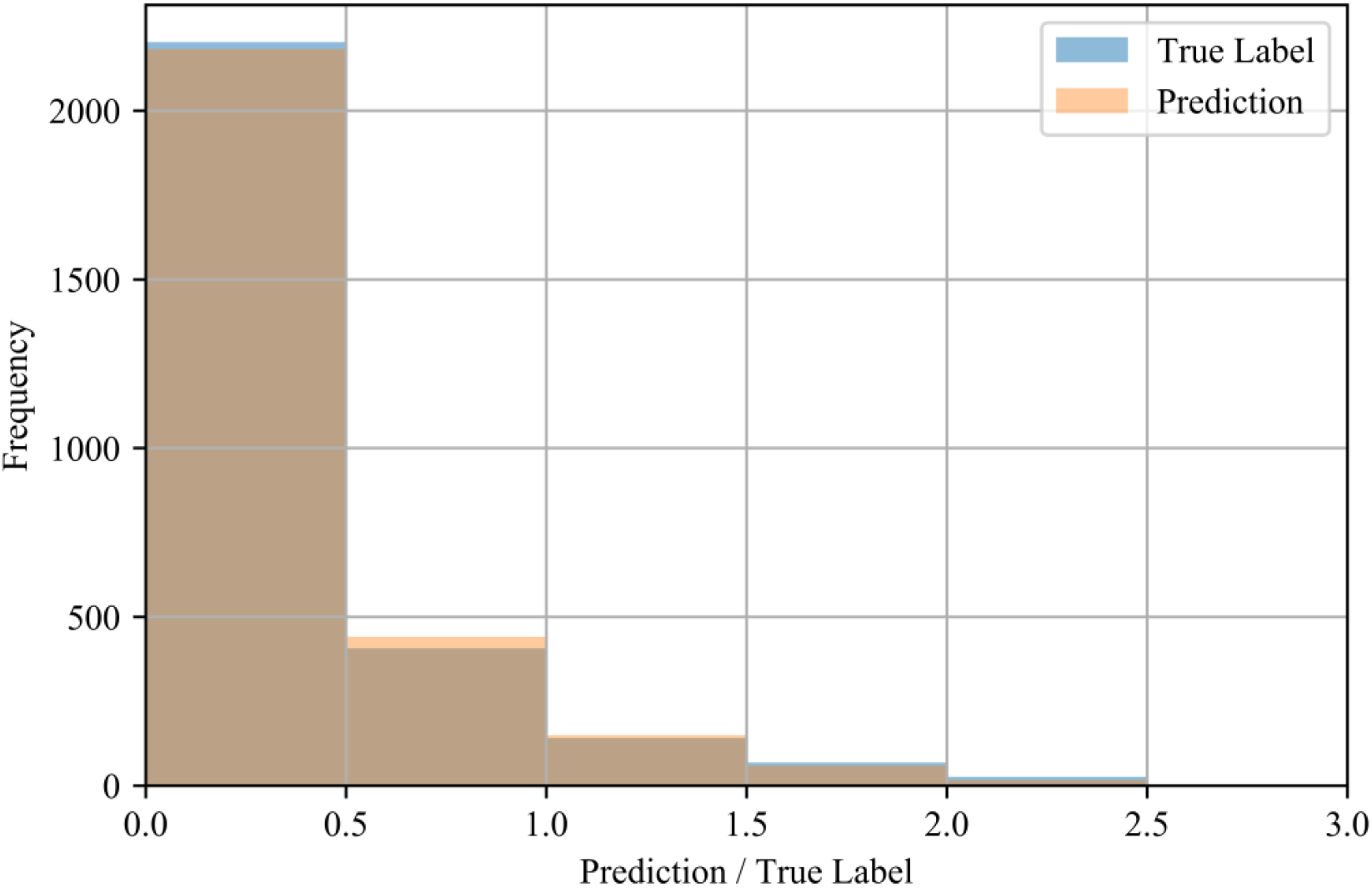
Distributions of test labels and predictions (*N* = 2847) for the interpolation validation method.

**Figure 5.**
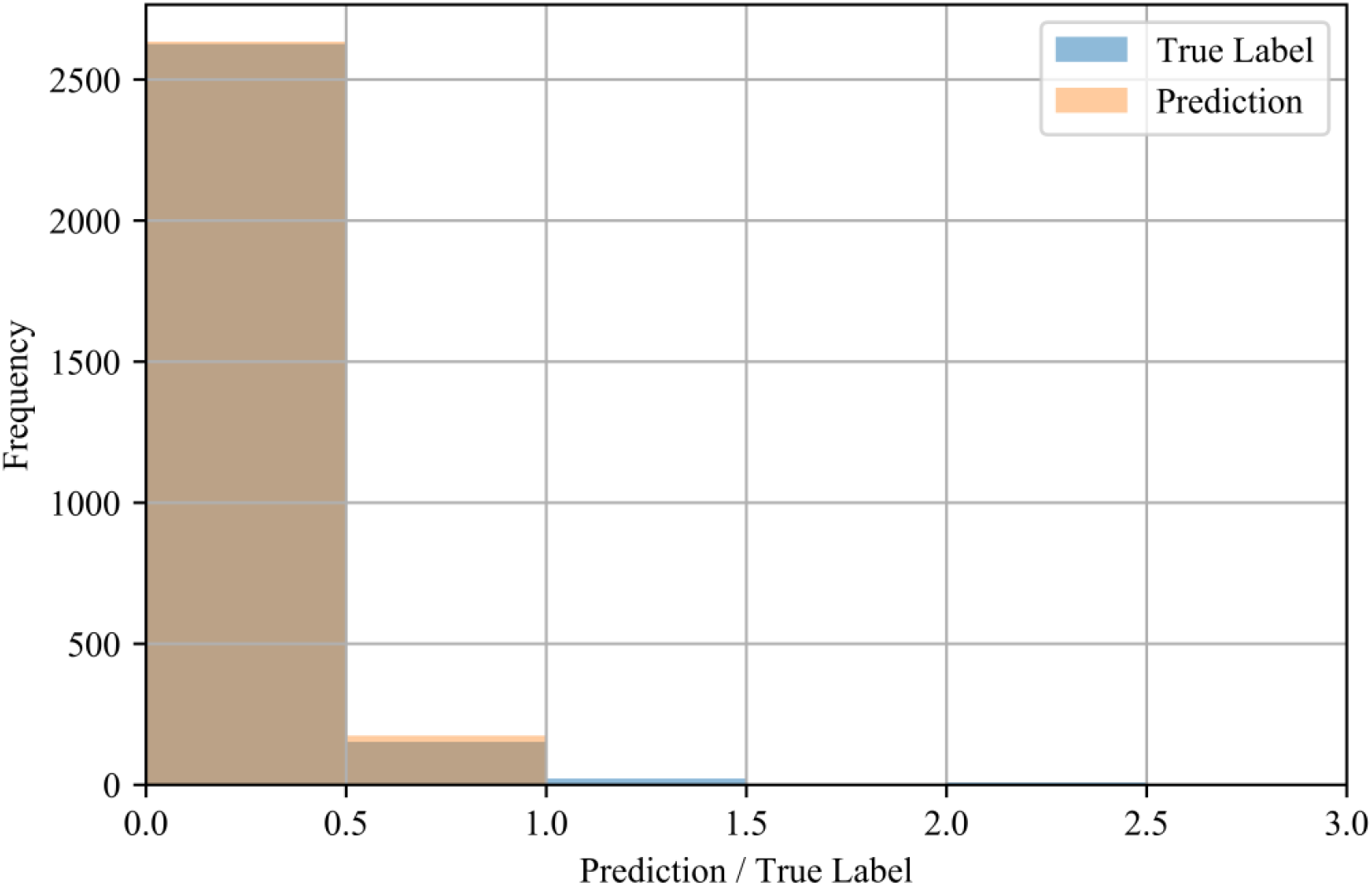
Distributions of test labels and predictions (*N* = 2811) for the extrapolation validation method.

**Figure 6.**
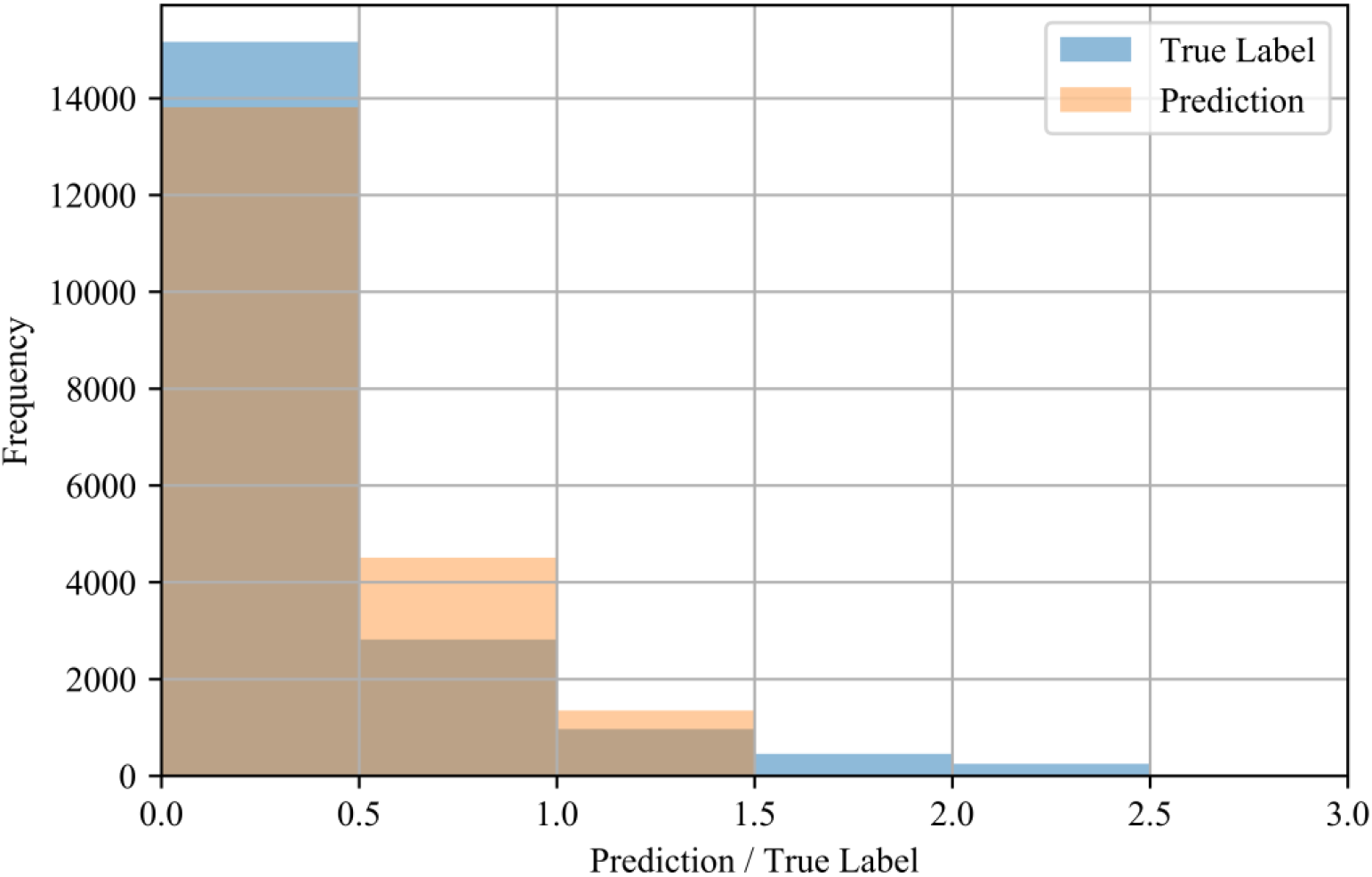
Distributions of test labels and predictions (*N* = 19,669) for the cross-validation method.

Further analysis shows that the performance of the models varies with the values of the labels. In both the in-distribution and cross-validation methods, the test MAE is lowest for samples with labels 0.0 (see Table 8 and Table 10), followed by label range 0.0 − 0.5. In the out-of-distribution method, the test MAE is lowest for samples with labels from 0.0 − 0.5 (see Table 9). For all validation methods, the mean MAE and median percent errors also increase with label bins greater 1.0, showing a decrease in accuracy for larger CIG.

**Table 8.** Test errors and median percent errors of label bins of size 0.5 for the in-distribution validation method.

| Upper Threshold | Count | Test Mean Non-Abs. Error | Test Mean Abs. Error (MAE) | Test Percent Error |
| --- | --- | --- | --- | --- |
| 0.0 | 20 | $0.000 \pm 0.000$ | $0.000 \pm 0.000$ | N/A |
| 0.5 | 2183 | $0.011 \pm 0.052$ | $0.017 \pm 0.050$ | 0.01 |
| 1.0 | 408 | $0.003 \pm 0.076$ | $0.047 \pm 0.060$ | 0.00 |
| 1.5 | 140 | $-0.052 \pm 0.139$ | $0.094 \pm 0.115$ | -0.02 |
| 2.0 | 68 | $-0.104 \pm 0.205$ | $0.158 \pm 0.167$ | -0.04 |
| 2.5 | 26 | $-0.283 \pm 0.309$ | $0.297 \pm 0.294$ | -0.08 |
| 3.0 | 2 | $-1.108 \pm 0.470$ | $1.108 \pm 0.470$ | -0.43 |

**Table 9.** Test errors and median percent errors of label bins of size 0.5 for the out-of-distribution validation method.

| Upper Threshold | Count | Test Mean Non-Abs. Error | Test Mean Abs. Error (MAE) | Test Percent Error |
| --- | --- | --- | --- | --- |
| 0.0 | 19 | $0.076 \pm 0.000$ | $0.076 \pm 0.000$ | N/A |
| 0.5 | 2607 | $0.034 \pm 0.096$ | $0.071 \pm 0.074$ | 0.44 |
| 1.0 | 152 | $-0.161 \pm 0.228$ | $0.225 \pm 0.164$ | -0.25 |
| 1.5 | 22 | $-0.648 \pm 0.222$ | $0.648 \pm 0.222$ | -0.52 |
| 2.0 | 3 | $-1.044 \pm 0.147$ | $1.044 \pm 0.147$ | -0.60 |
| 2.5 | 8 | $-1.464 \pm 0.116$ | $1.464 \pm 0.116$ | -0.67 |

**Table 10.** Test errors and median percent errors of label bins of size 0.5 for the cross-validation method.

| Upper Threshold | Count | Test Mean Non-Abs. Error | Test Mean Abs. Error (MAE) | Test Percent Error |
| --- | --- | --- | --- | --- |
| 0.0 | 114 | $-0.059 \pm 0.086$ | $0.059 \pm 0.086$ | N/A |
| 0.5 | 15,056 | $-0.073 \pm 0.174$ | $0.109 \pm 0.153$ | 0.493 |
| 1.0 | 2815 | $-0.010 \pm 0.282$ | $0.217 \pm 0.181$ | -0.006 |
| 1.5 | 960 | $0.333 \pm 0.337$ | $0.393 \pm 0.265$ | -0.299 |
| 2.0 | 451 | $0.719 \pm 0.370$ | $0.719 \pm 0.370$ | -0.391 |
| 2.5 | 246 | $1.141 \pm 0.321$ | $1.141 \pm 0.321$ | -0.459 |
| 3.0 | 27 | $1.362 \pm 0.266$ | $1.225 \pm 0.266$ | -0.486 |

## Discussion

### Principal Results

Our results suggest that traditional, non-time series machine learning models can predict future CIG to an appreciable degree of accuracy, as suggested by the high values and strong linear calibration relationships between the labels and predictions in all validation methods.

A comparison of our results for all validation methods suggests differences in the predictive performance of machine learning models across the varying use cases. The in-distribution method has the lowest test mean error and median percent error, which is to be expected as the test samples are obtained from the same distribution as the training samples. Intuitively, while samples in the in-distribution method are unordered (i.e., no temporal features are included), the availability of samples across the entire temporal range in the training set allows the validation and test samples to interpolate between these training samples.

The out-of-distribution method achieves a test mean error which is higher than that of the in-distribution method. This is expected, as evolving COVID-19 infection trajectories observed most countries give distributions of training samples from earlier dates that may differ greatly from those of validation and test samples from later dates (i.e., data shift), which often leads to poor generalization (e.g., over-fitting) of machine learning models.

Conversely, even though the cross-validation method contains the training and validation sets within the same date range, the cross-validation method also separates countries across these sets (i.e., the 10 folds), such that there is no country which has samples in both the training and validation sets. This difference leads to higher test mean errors and median percent errors than both other methods, which suggests that including training samples from the same country as the validation samples is more important than ensuring temporal overlap. We speculate that this occurs because the unique cultural dimensions per country may potentially act as a categorical rather than continuous features, for each country. In such cases, the cultural dimensions observed in the training set would be considered irrelevant to cultural dimensions within the validation set.

Performance also varies depending on the value of the label (see Table 8, Table 9, and Table 10), which may be due to the imbalanced frequency of training samples. That is, the rareness of samples with higher CIG compared to lower CIG in the training set may be the cause of their comparatively poorer performance.

In Figure 2 and Figure 3, we also observe constraints of the trained AdaBoost regression models. Discretization of the prediction values may be due to the low number of estimators used in the lowest mean test error configuration, as seen in Table 3. The low number of estimators in these configurations may also restrict the predictions to a maximum of 1.5 selected to the relatively lower number of samples with labels greater than 1.5 (see Figure 5 and Figure 6). Label ranges with the most samples are selected over underrepresented ranges as candidates for prediction values in the discretized AdaBoost regression models. While additional estimators in the AdaBoost regression models may result in less discrete prediction values, they may also cause over-fitting by increasing the complexity of the models.

### Limitations

First, the scores in the OxCGRT and Hofstede’s cultural dimensions datasets are imprecise. NPI enforcement levels and definitions may vary even between countries with the same scores, while countries sharing similar cultural dimension scores may have unobserved differences in terms of cultural practices. Second, by predicting the CIG 14 days in advance of the current date, the models do not account for information regarding changes in NPIs between the current date and the date-to-predict. Third, the CIG is a measure of the change in the cumulative number of confirmed infections and may not necessarily be correlated with the change in the daily number of confirmed infections nor the actual transmission rate of COVID-19. For example, changing biased and/or unreliable testing policies (e.g., prioritizing high-risk patients) may lead to a misleading representation of the infection growth within the general population.

## Conclusion

In this study, we train five non-time series machine learning models to predict the CIG 14 days into the future, using NPI features extracted from the OxGCRT dataset [1], as well as cultural norm features extracted from Hofstede’s cultural dimensions [34]. Together, these features enable the prediction of near-future CIG in multiple machine learning models. Specifically, we observe that random forest regression and AdaBoost regression result in the most accurate predictions out of the five evaluated machine learning models.

We observe differences in the predictive performance of the machine learning models across the three validation methods, with the highest accuracy with the in-distribution method and the lowest with the cross-validation method. These differences in performance suggest that the models have varying levels of accuracy depending on use case. Specifically, predictions are expected to have higher accuracies when existing data from the same country in nearby dates are available (i.e., in-distribution method). This enables use cases such as predicting the CIG over the upcoming 14 days from the current date. The decrease in accuracy when data from nearby dates are unavailable (i.e., out-of-distribution method) suggest weaker performance in predicting the CIG over 14 days for relatively distanced future dates. We observe the greatest decrease in performance when data from the same country is unavailable (i.e., cross-validation method). However, with all validation methods, we observe appreciable calibration measures between the predictions and labels of the test set.

Due to the rapidly growing body of work related to predicting COVID-19 infection rates, one cannot claim state-of-the-art results. However, this work provides new considerations in the use of NPIs and cultural dimensions for predicting the national growth of confirmed infections of the COVID-19 pandemic and other infectious diseases using non-time series machine learning models. These experiments also provide insight into validation methods for different applications of the models. As the availability of such data increases and the nature of the data continues to evolve, we expect that simple and straightforward models such as these may prove to be more accurate and generalizable, improving overall predictive performance.

## Data Availability

All data used in this manuscript is open-sourced and readily accessible online.

## Acknowledgments

Frank Rudzicz is supported by a CIFAR Chair in Artificial Intelligence.

## References

1. Hale T, Petherick A, Phillips T, Webster S. Variation in government responses to COVID-19. Blavatnik Sch Gov Work Pap. 2020;31.

2. Flaxman S, Mishra S, Gandy A, Unwin HJT, Coupland H, Mellan TA, et al. Estimating the number of infections and the impact of non-pharmaceutical interventions on COVID-19 in European countries: technical description update. arXiv Prepr 200411342. 2020;

3. McCoy LG, Smith J, Anchuri K, Berry I, Pineda J, Harish V, et al. CAN-NPI: A Curated Open Dataset of Canadian Non-Pharmaceutical Interventions in Response to the Global COVID-19 Pandemic. medRxiv. Cold Spring Harbor Laboratory Press; 2020;

4. Hens N, Vranck P, Molenberghs G. The COVID-19 epidemic, its mortality, and the role of non-pharmaceutical interventions. Eur Hear J Acute Cardiovasc Care. SAGE Publications Sage UK: London, England; 2020;9(3):204–208.

5. Chang SL, Harding N, Zachreson C, Cliff OM, Prokopenko M. Modelling transmission and control of the COVID-19 pandemic in Australia. arXiv Prepr 200310218. 2020;

6. Davies NG, Kucharski AJ, Eggo RM, Gimma A, Edmunds WJ, Jombart T, et al. Effects of non-pharmaceutical interventions on COVID-19 cases, deaths, and demand for hospital services in the UK: a modelling study. Lancet Public Heal. Elsevier; 2020;

7. Cowling BJ, Ali ST, Ng TWY, Tsang TK, Li JCM, Fong MW, et al. Impact assessment of non-pharmaceutical interventions against coronavirus disease 2019 and influenza in Hong Kong: an observational study. Lancet Public Heal. Elsevier; 2020;

8. Pan A, Liu L, Wang C, Guo H, Hao X, Wang Q, et al. Association of public health interventions with the epidemiology of the COVID-19 outbreak in Wuhan, China. Jama. American Medical Association; 2020;323(19):1915–1923.

9. Brauner JM, Sharma M, Mindermann S, Stephenson AB, Gavenčiak T, Johnston D, et al. The effectiveness and perceived burden of nonpharmaceutical interventions against COVID-19 transmission: a modelling study with 41 countries. medRxiv. Cold Spring Harbor Laboratory Press; 2020;

10. Aledort JE, Lurie N, Wasserman J, Bozzette SA. Non-pharmaceutical public health interventions for pandemic influenza: an evaluation of the evidence base. BMC Public Health. Springer; 2007;7(1):208.

11. Merler S, Ajelli M, Fumanelli L, Gomes MFC,y Piontti AP, Rossi L, et al. Spatiotemporal spread of the 2014 outbreak of Ebola virus disease in Liberia and the effectiveness of non-pharmaceutical interventions: a computational modelling analysis. Lancet Infect Dis. Elsevier; 2015;15(2):204–211.

12. Cowling BJ, Fung ROP, Cheng CKY, Fang VJ, Chan KH, Seto WH, et al. Preliminary findings of a randomized trial of non-pharmaceutical interventions to prevent influenza transmission in households. PLoS One. Public Library of Science; 2008;3(5):e2101.

13. Ferguson N, Laydon D, Nedjati-Gilani G, Imai N, Ainslie K, Baguelin M, et al. Report 9: Impact of non-pharmaceutical interventions (NPIs) to reduce COVID19 mortality and healthcare demand. Imp Coll London. 2020;10:77482.

14. Lai S, Ruktanonchai NW, Zhou L, Prosper O, Luo W, Floyd JR, et al. Effect of non-pharmaceutical interventions for containing the COVID-19 outbreak in China. medRxiv. Cold Spring Harbor Laboratory Preprints; 2020;

15. Delamater PL, Street EJ, Leslie TF, Yang YT, Jacobsen KH. Complexity of the basic reproduction number (R0). Emerg Infect Dis. Centers for Disease Control and Prevention; 2019;25(1):1.

16. Bjørnstad ON, Finkenstädt BF, Grenfell BT. Dynamics of measles epidemics: estimating scaling of transmission rates using a time series SIR model. Ecol Monogr. Wiley Online Library; 2002;72(2):169–184.

17. Kermack WO, McKendrick AG. A contribution to the mathematical theory of epidemics. Proc R Soc london Ser A, Contain Pap a Math Phys character. The Royal Society London; 1927;115(772):700–721.

18. Nåsell I. The quasi-stationary distribution of the closed endemic SIS model. Adv Appl Probab. JSTOR; 1996;895–932.

19. Li MY, Muldowney JS. Global stability for the SEIR model in epidemiology. Math Biosci. Elsevier; 1995;125(2):155–164.

20. Abrams D, Grant PR. Testing the social identity relative deprivation (SIRD) model of social change: The political rise of Scottish nationalism. Br J Soc Psychol. Wiley Online Library; 2012;51(4):674–689.

21. Chen Y-C, Lu P-E, Chang C-S, Liu T-H. A Time-dependent SIR model for COVID-19 with undetectable infected persons. IEEE Trans Netw Sci Eng. IEEE; 2020;

22. Calafiore GC, Novara C, Possieri C. A modified sir model for the covid-19 contagion in italy. arXiv Prepr 200314391. 2020;

23. Yang Z, Zeng Z, Wang K, Wong S-S, Liang W, Zanin M, et al. Modified SEIR and AI prediction of the epidemics trend of COVID-19 in China under public health interventions. J Thorac Dis. AME Publications; 2020;12(3):165.

24. Fernández-Villaverde J, Jones CI. Estimating and Simulating a SIRD Model of COVID-19 for Many Countries, States, and Cities. 2020.

25. Liu D, Clemente L, Poirier C, Ding X, Chinazzi M, Davis JT, et al. A machine learning methodology for real-time forecasting of the 2019-2020 COVID-19 outbreak using Internet searches, news alerts, and estimates from mechanistic models. arXiv Prepr 200404019. 2020;

26. Pinter G, Felde I, Mosavi A, Ghamisi P, Gloaguen R. COVID-19 Pandemic Prediction for Hungary; a Hybrid Machine Learning Approach. Mathematics. Multidisciplinary Digital Publishing Institute; 2020;8(6):890.

27. Alimadadi A, Aryal S, Manandhar I, Munroe PB, Joe B, Cheng X. Artificial intelligence and machine learning to fight COVID-19. American Physiological Society Bethesda, MD; 2020.

28. Yan L, Zhang H-T, Xiao Y, Wang M, Sun C, Liang J, et al. Prediction of criticality in patients with severe Covid-19 infection using three clinical features: a machine learning-based prognostic model with clinical data in Wuhan. MedRxiv. Cold Spring Harbor Laboratory Press; 2020;

29. Randhawa GS, Soltysiak MPM, El Roz H, de Souza CPE, Hill KA, Kari L. Machine learning using intrinsic genomic signatures for rapid classification of novel pathogens: COVID-19 case study. PLoS One. Public Library of Science San Francisco, CA USA; 2020;15(4):e0232391.

30. Harmon SA, Sanford TH, Xu S, Turkbey EB, Roth H, Xu Z, et al. Artificial intelligence for the detection of COVID-19 pneumonia on chest CT using multinational datasets. Nat Commun. Nature Publishing Group; 2020;11(1):1–7.

31. Van Bavel JJ, Baicker K, Boggio PS, Capraro V, Cichocka A, Cikara M, et al. Using social and behavioural science to support COVID-19 pandemic response. Nat Hum Behav. Nature Publishing Group; 2020;1–12.

32. Zhu N, O J, Lu HJ, Chang L. Debate: Facing uncertainty with (out) a sense of control--cultural influence on adolescents’ response to the COVID-19 pandemic. Child Adolesc Ment Health. Wiley Online Library; 2020;25(3):173–174.

33. Dryhurst S, Schneider CR, Kerr J, Freeman ALJ, Recchia G, Van Der Bles AM, et al. Risk perceptions of COVID-19 around the world. J Risk Res. Taylor & Francis; 2020;1–13.

34. Hofstede G, Bond MH. Hofstede’s culture dimensions: An independent validation using Rokeach’s value survey. J Cross Cult Psychol. Sage Publications Sage CA: Thousand Oaks, CA; 1984;15(4):417–433.

35. McKinney W, others. pandas: a foundational Python library for data analysis and statistics. Python High Perform Sci Comput. Seattle; 2011;14(9).

36. Pedregosa F, Varoquaux G, Gramfort A, Michel V, Thirion B, Grisel O, et al. Scikit-learn: Machine learning in Python. J Mach Learn Res. JMLR. org; 2011;12:2825–2830.

37. Hofstede G. Dimension data matrix [Internet]. GeertHoftstede.com. 2015. https://geerthofstede.com/research-and-vsm/dimension-data-matrix/

38. Soares AM, Farhangmehr M, Shoham A. Hofstede’s dimensions of culture in international marketing studies. J Bus Res. Elsevier; 2007;60(3):277–284.

39. Hofstede G. Dimensionalizing cultures: The Hofstede model in context. Online readings Psychol Cult. 2011;2(1):919–2307.

40. Dong E, Du H, Gardner L. An interactive web-based dashboard to track COVID-19 in real time. Lancet Infect Dis. Elsevier; 2020;20(5):533–534.

41. Ross BC. Mutual information between discrete and continuous data sets. PLoS One. Public Library of Science; 2014;9(2):e87357.

42. Pirbazari AM, Chakravorty A, Rong C. Evaluating feature selection methods for short-term load forecasting. 2019 IEEE Int Conf Big Data Smart Comput. 2019. p. 1–8.

43. Hoerl AE, Kennard RW. Ridge regression: Biased estimation for nonorthogonal problems. Technometrics. Taylor & Francis Group; 1970;12(1):55–67.

44. Breiman L. Random forests. Mach Learn. Springer; 2001;45(1):5–32.

45. Freund Y, Schapire RE. A decision-theoretic generalization of on-line learning and an application to boosting. J Comput Syst Sci. Elsevier; 1997;55(1):119–139.

46. Drucker H, Burges CJ, Kaufman L, Smola A, Vapnik V. Support vector regression machines. Adv Neural Inf Process Syst. 1996;9:155–161.

47. Willmott CJ, Matsuura K. Advantages of the mean absolute error (MAE) over the root mean square error (RMSE) in assessing average model performance. Clim Res. 2005;30(1):79–82.

48. Steyerberg EW, Vickers AJ, Cook NR, Gerds T, Gonen M, Obuchowski N, et al. Assessing the performance of prediction models: a framework for some traditional and novel measures. Epidemiology. NIH Public Access; 2010;21(1):128.

49. Schaffer C. Selecting a classification method by cross-validation. Mach Learn. Springer; 1993;13(1):135–143.

50. Browne MW. Cross-validation methods. J Math Psychol. Elsevier; 2000;44(1):108–132.

51. Ardabili SF, Mosavi A, Ghamisi P, Ferdinand F, Varkonyi-Koczy AR, Reuter U, et al. Covid-19 outbreak prediction with machine learning. Available SSRN 3580188. 2020;

